# An Exploratory Study of Host Plasma Proteomic Signatures that Distinguish Active Syphilis in Adults

**DOI:** 10.64898/2026.03.04.26347505

**Authors:** Cody Chou, Shad R. Morton, Kelika A Konda, Silver Vargas, Michael Reyes-Diaz, Francesca Vasquez, Carlos F. Caceres, Jeffrey D. Klausner, Trey Toombs, Rushdy Ahmad, Lao-Tzu Allan-Blitz

## Abstract

Syphilis remains a major public health concern. However, current serologic assays are limited in their ability to distinguish active from previously treated disease. We applied tandem mass tag-based quantitative proteomics to plasma from 10 adults with active syphilis and 10 age- and gender-matched non-diseased controls. We identified 54 differentially regulated proteins (36 upregulated, 18 downregulated). Those proteins map to immune and inflammatory responses, acute-phase signaling, coagulation and vascular pathways, and cellular stress processes. Three sets of between 2-5 proteins achieved ≥99% discrimination between cases and controls. Our exploratory findings support proteomics as a potential tool to develop novel syphilis diagnostics.

## Background

In 2020, more than 8 million people worldwide had syphilis, a sexually transmitted disease caused by *Treponema pallidum* (1). Syphilis is diagnosed using serologic tests that detect antibodies against *T. pallidum* (treponemal) and lipoidal (non-treponemal) antigens (2). However, treponemal antibodies do not readily distinguish active from previously treated infection. Lipoidal test titers indicate disease activity and response to treatment; however, they lack specificity and may yield false-positive results in patients with other medical conditions (2). They also must be repeated over time in some instances, during which patients may be lost to follow-up.

Recent work has identified differences in cytokine expression between individuals with active syphilis and non-diseased controls. Those cytokine signatures include elevated pro-inflammatory chemokines, Th1-type cytokines, pro-inflammatory interleukins, and tumor necrosis factor (TNF) family mediators (3, 4, 5). Distinct protein patterns could support the development of novel diagnostic assays. We evaluated plasma protein profiles to identify additional host-response biomarkers that could distinguish individuals with active syphilis from those without the disease.

## Methods

### Plasma samples

We used plasma samples previously collected from patients with and without syphilis as part of ongoing studies in Peru (6). We defined active syphilis as a positive treponemal test and a lipoidal test titer ≥ 1:4 with compatible clinical symptoms consistent with secondary syphilis when present. We selected plasma samples matched on age, gender, between syphilis cases and controls (samples from individuals with negative treponemal and lipoidal test results). We only included samples from people without HIV.

### Mass Spectrometry

We used tandem mass tag labeling to quantify peptide abundances across samples within a single mass spectrometry run (7). We depleted the 14 most abundant plasma proteins using Pierce Top 14 columns (Thermo Fisher Scientific, United States), then reduced the remaining proteins with tris (2-carboxyethyl) phosphine, alkylated with iodoacetamide, and digested overnight with trypsin. We then labeled the resulting peptides with the TMTpro 18-plex kit (Thermo Fisher Scientific), pooled and dried, and fractionated them into 20 fractions using high-pH reversed-phase chromatography prior to mass spectrometry analysis.

We processed peptide spectra in Proteome Discoverer (Thermo Fisher Scientific) and used the Chimerys database to identify and quantify proteins. We normalized protein abundances to a pseudo-reference channel defined as the mean across samples. We imputed missing values within diagnostic groups using k-nearest neighbors and excluded proteins with more than 50% missingness. We assessed differential expression using limma in R version 4.5.0 (R Foundation, Austria) and performed feature selection and classification modeling in Python version 1.7 (Python Software Foundation, United States).

We used the STRING database of known and predicted protein-protein associations for gene ontology and functional enrichment analyses. To characterize global expression patterns, we generated relative log expression plots and performed principal component analysis to assess separation between cases and controls, as well as heterogeneity between secondary and early latent syphilis. We then examined the functional significance of the observed proteomic alterations through protein–protein interaction network analysis.

We evaluated a subset of the proteins with differential concentrations which could serve as effective biomarkers using a recursive feature elimination with cross-validation. That involved training four machine-learning classification models (linear support vector machines, logistic regression, decision tree, and random forest) on the data while iteratively removing the least important feature and evaluating performance at each step. We calculated the area under the resulting receiver operating characteristics curves (AUC) and plotted confusion matrices using cross-validated predictions (scikit-learn) across eleven candidate models: logistic regression, support vector machine, decision tree, random forest, naïve Bayes, k-nearest neighbors, linear discriminant analysis, gradient boosting, extra trees, bagging, and a soft-voting ensemble (logistic regression, decision tree, and support vector machine). We then selected the final model based on mean accuracy and lowest standard deviation across 1,000 permutation tests.

### Ethical Considerations

The Universidad Cayetano Heredia Institutional Review Board (CIEI #103093) approved the secondary use of stored samples. The Mass General Brigham Institutional Review Board determined that the analysis of de-identified data generated from those samples did not constitute human subjects’ research.

## Results

We analyzed 10 case samples (5 secondary syphilis and 5 early latent syphilis) and 10 controls. All samples were obtained from adults (80% male) between 2019 and 2022.

Samples from controls exhibited upshifted median log10-relative peptide intensities of 0.899. Samples from cases with early latent syphilis exhibited median log10-relative peptide intensities values of 0.052, while samples from cases with secondary syphilis exhibited downshifted median log10-relative peptide intensities of −0.699 (Figure 1a). Principal component analyses demonstrated clear separation between syphilis cases and controls. The first principal component explained 35.7% of the total variance and separated controls from syphilis cases. The second principal component explained an additional 16.4% and distinguished secondary syphilis from early latent syphilis. Together, those accounted for 52.1% of the total variability (Figure 1b).

**Figure 1.**
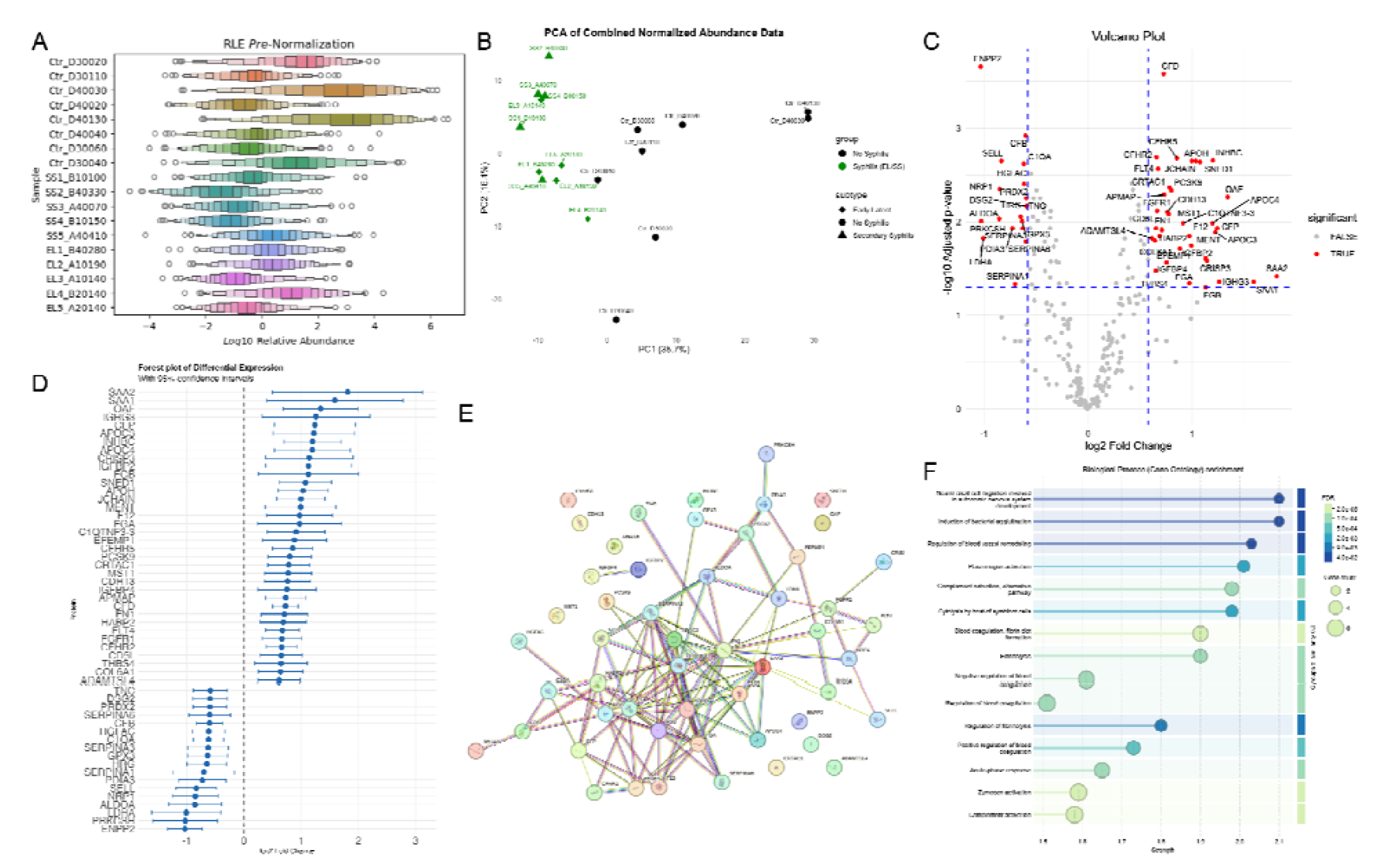
Primary Proteomic Differences Between 10 Cases of Syphilis and 10 Non-Diseased Controls. (A) Relative Log Expression for all samples show heterogeneity in the control group (top 8), lower expression for secondary syphilis (middle) with early latent (bottom) slightly higher (RLE = Relative Log Expression). (B) Principal Component Analysis captures the variability between the control and syphilis group, and the heterogeneity of the secondary and early latent groups (Ctr = Control, SS = Secondary Syphilis, EL = Early latent Syphilis). (C) Volcano plot showing significantly up and down regulated proteins; differences were considered significant when p-value < 0.05 (1.3 - log10) and log2 fold change > absolute value 0.58 (1.5-fold change). Case = Syphilis, Control = Non-Diseased. (D) Forest plot with 95% confidence interval of differentially expressed proteins ordered by fold change. (E) Protein-protein interactions found by STRING database. (F) Top-15 biological processes by strength shown with False Discovery Rate and protein count.

We identified 54 proteins that differed significantly between syphilis cases and controls (p<0.05; >1.5-fold change), including 36 upregulated and 18 downregulated among cases (Figure 1c and 1d). Serum Amyloid A2 (SAA2) (1.81 log2 fold change; 95% CI: 0.50–3.13), Serum Amyloid A1 (SAA1) (1.59 log2 fold change; 95% CI: 0.40– 2.78), and Out At First (1.34 log2 fold change; 95% CI: 0.69–2.00) ranked among the most upregulated proteins. Lactate Dehydrogenase A (−1.01 log2 fold change; 95% CI: −1.61 to −0.40), Protein Kinase C Substrate 80K-H (PRKCSH) (−1.03 log2 fold change; 95% CI: −1.59 to −0.46), and Ectonucleotide Pyrophosphatase/Phosphodiesterase 2 (ENPP2) (−1.03 log2 fold change; 95% CI: −1.34 to −0.73) ranked among the most downregulated proteins. These upregulated and downregulated proteins show strong protein-protein interactions found by STRING database (Figure 1e). Figure 1f shows the gene ontology associations, while the Supplemental Table shows related functions of known protein signatures.

Recursive feature elimination with cross-validation produced a ranked feature list from the 54 candidate proteins, from which 9 candidate proteins were retained for panel evaluation (Figure 2a-b). Confusion matrices reflected 18 samples since tandem mass tag labeling limits multiplexing capacity to 18 per batch. Three candidate biomarker panels were evaluated. Figure 2a summarizes the performance metrics for the selected models, and Figure 2c-e shows the corresponding confusion matrices.

**Figure 2.**
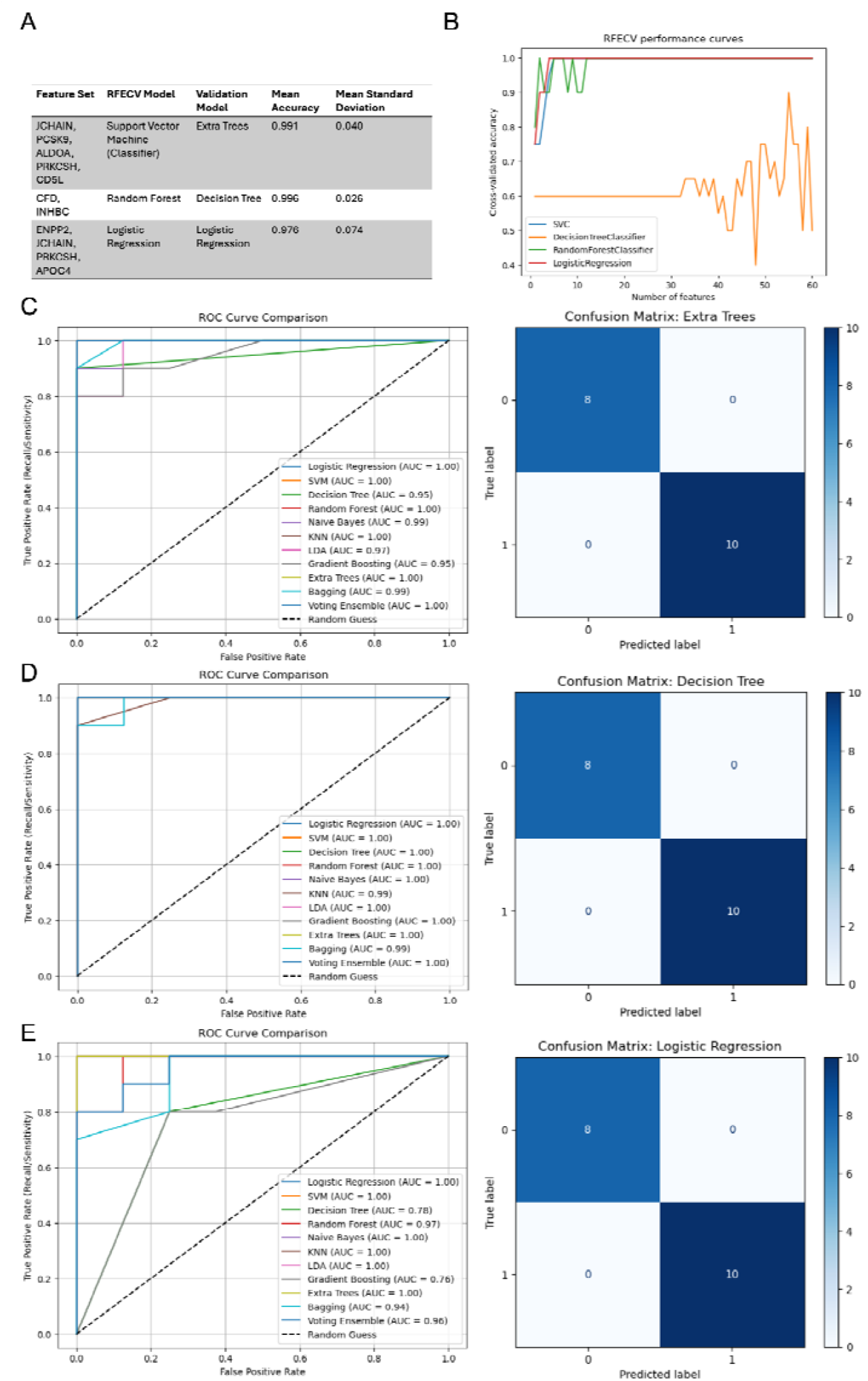
Identification of Candidate Protein Signatures using 11 Models with Corresponding AUC Plots and Confusion Matrices. (A) Candidate proteins chosen by Recursive Feature Elimination with Cross-Validation with permutation scores (B) Recurrent feature elimination with cross-validation starting with all 54 features (+6 isoforms) (right), cross-validating by accuracy and eliminating one feature until none remain. (C) Receiver operating characteristics curve and confusion matrix for first candidate protein panel (JCHAIN, PCSK9, ALDOA, PRKCSH, CD5L). (D) Receiver operating characteristics curve and confusion matrix for second candidate protein panel (CFD, INHBC). (E) Receiver operating characteristics curve and confusion matrix for third candidate protein panel (ENPP2, JCHAIN, PRKCSH, APOC4).

The five-protein panel (JCHAIN, PCSK9, ALDOA, PRKCSH, CD5L; Figure 2c) achieved an AUC of 1.00 (95% CI: 1.00–1.00) using an Extra Trees classifier. The two-protein panel (CFD, INHBC; Figure 2d) achieved an AUC of 0.996 (95% CI: 0.90–1.00) using a Decision Tree classifier. The four-protein panel (ENPP2, JCHAIN, PRKCSH, APOC4; Figure 2e) achieved an AUC of 1.00 (95% CI: 1.00–1.00) using Logistic Regression.

## Discussion

In a pilot evaluation, protein signatures differed significantly between cases of active syphilis and controls. The 9 proteins most predictive of active syphilis originated from 1) immune and inflammatory responses (JCHAIN, CD5L, CFD, Serum Amyloid A1 and 2, and INHBC), 2) coagulation and vascular biology (PCSK9, APOC4, and ENPP2), and 3) cellular stress and damage responses (ALDOA and PRKCSH). Those pathways align with known features of *T. pallidum* infection and support the biological relevance of the proteomic shifts we observed (8, 9). Prior studies of host responses to *T. pallidum* similarly identified inflammatory and vascular pathways, including elevations in TNF-family cytokines, IL-6, IL-1β, and vascular endothelial growth factor-related mediators in active syphilis (3, 4). In addition to immune activation, infection may also trigger vascular responses (9). Our study identified increased levels of FLT4, which encodes vascular endothelial growth factor receptor 3, which is typically expressed on lymphatic endothelium (10). We also detected up-regulation of extracellular matrix and adhesion proteins such as thrombospondin-4 (11).

We identified several upregulated proteins that warrant closer examination. JCHAIN transports immunoglobulins across mucosal epithelia and plays a role in humoral immunity (12). SAA1 participates in host defense and inflammation by acting as an opsonin for certain bacteria, chemoattracting immune cells, and inducing enzymes that degrade the extracellular matrix (13). During inflammation, SAA2 and SAA1 can increase by 10^3^-fold and function as cytokine-like mediators (14).

Our analysis found that several proteins were downregulated. PRKCSH (protein kinase C substrate 80K-H) is overexpressed in most tumors, and its suppression has been associated with enhanced anti-tumor immunity. Lower PRKCSH levels may allow stronger immune surveillance by impairing the maturation of regulatory glycoproteins on tumor or immune cells (15). In the context of infection, reduced PRKCSH expression might alter the presentation of glycosylated antigens. Finally, despite the small sample size, our results identified promising proteomic signatures that may distinguish active syphilis cases from controls.

## Limitations

Our study had several limitations. First, the small sample size limits the precision of our findings, and the machine-learning analysis may render the model susceptible to overfitting despite cross-validation. The classification results should be interpreted as hypothesis-generating. Further, because we evaluated thousands of proteins, some of the observed associations may reflect type I error. Despite those limitations, our results indicate that proteomic analyses are promising strategies for furthering our understanding of the pathophysiology of *T. pallidum* infection and detection.

## Conclusion

In this pilot study, host plasma proteomic profiles distinguished adults with active syphilis from non-diseased controls. Although the sample size was small, our findings support further investigation of host-response proteomic biomarkers to advance our understanding of the disease and to evaluate their potential as tools to improve syphilis diagnosis.

## Author Contributions

Conception and design of the study: R.A. and L.A.B. Data acquisition and curation: S.R.M., K.A.K., M.R.D., F.V., C.F.C, S.V., T.T., R.A and L.A.B. Formal analysis and interpretation of data: C.C., S.R.M., K.A.K., M.R.D., F.V., C.F.C, S.V., J.D.K., T.T., R.A., and L.A.B. Manuscript drafting: C.C., and L.A.B. Manuscript review and editing: C.C., S.R.M., K.A.K., M.R.D., F.V., C.F.C, S.V., J.D.K., T.T., R.A., and L.A.B. Supervision of the study: L.A.B. Funding acquisition: J.D.K., R.A., and L.A.B. All authors contributed to the article and approved the submitted version.

## Data Availability Statement

The data underlying this article will be shared on reasonable request to the corresponding author.

