## Supplemental Table for "An Exploratory Study of Host Plasma Proteomic Signatures that Distinguish Active Syphilis in Adults"

### Supplementary Table: Biological Characterization of Up-Regulated and Down-Regulated Proteins.

| Up-Regulated Protein (Gene Name) | Log$_2$ Fold Change | Characterization of protein | Supplemental Citations |
|---|---|---|---|
| SAA2 | 1.81 | SAA2 is an acute-phase apolipoprotein produced primarily by hepatocytes in response to IL-1, IL-6, and TNF through NF-κB/STAT3 signaling. During inflammation, SAA2 (together with SAA1) can increase up to $10^3$-fold and functions as a cytokine-like mediator. It signals through TLR2, TLR4, and FPR2 to activate NF-κB, induce chemokine expression, and promote neutrophil and monocyte chemotaxis. At barrier sites, SAA2 also drives pathogenic Th17 cell differentiation. | 1, 2, 3 |
| SAA1 | 1.59 | SAA1 is an acute-phase protein produced by the liver during infection and tissue injury. It promotes leukocyte chemotaxis and Th17 polarization through FPR2 and TLR2/TLR4 signaling. | 4 |
| OAF | 1.34 | OAF is a member of the BRICHOS-domain protein family. BRICHOS domains function as intramolecular chaperones that prevent amyloid-like protein aggregation. | 5 |
| IGHG3 | 1.26 | IGHG3 encodes the constant region of the IgG3 heavy chain. Compared to other IgG subclasses, IgG3 has a relatively long hinge region that allows greater Fab arm flexibility. It is effective at complement activation, Fcγ receptor binding, | 6, 7 |

| | | | |
|---|---|---|---|
| | | opsonization, and pathogen clearance. IGHG3 is also highly polymorphic with numerous G3m allotypes contributing to considerable genetic diversity across individuals. | |
| CFP | 1.24 | CFP encodes properdin, which binds and stabilizes the alternative-pathway C3 convertase (C3bBb). It amplifies C3b deposition and downstream inflammation. Properdin can also act as a pattern-recognition molecule released locally by leukocytes to initiate alternative-pathway activity on microbial and apoptotic cell surfaces. | 8, 9 |
| APOC3 | 1.22 | APOC3 is an apolipoprotein produced by the liver and intestine that associates with triglyceride-rich lipoproteins. It inhibits lipoprotein lipase and hepatic uptake of triglyceride-rich lipoproteins. Beyond lipid metabolism, APOC3 can act as an endogenous damage-associated molecular pattern (DAMP) that triggers inflammation by activating the NLRP3 inflammasome in human monocytes through TLR2/4 and Syk/caspase-8 signaling which leads to IL-1β release and monocyte activation. | 10, 11 |
| INHBC | 1.2 | INHBC encodes a member of the TGF-β superfamily. Can form homodimers or heterodimers (with other beta-subunits) to function as activins/inhibins. Expression data shows that INHBC is expressed in immune-relevant cell types (e.g., monocytes & neutrophils) and lymphoid/bone-marrow tissues | 12, 13 |

| | | | |
|---|---|---|---|
| APOC4 | 1.2 | APOC4 belongs to the apolipoprotein C family, which regulates triglyceride-rich lipoprotein metabolism. It influences lipoprotein lipase activity and hepatic uptake of lipoproteins. | 14 |
| CRISP3 | 1.14 | CRISP3 is a secreted glycoprotein belonging to the CAP superfamily. CRISP3 is upregulated in sepsis, severe dengue, HCV infection, and neutrophil-activation gene clusters identified in long COVID. It is thought to contribute to neutrophil degranulation and modulation of inflammation. | 15, 16 |
| IGFBP2 | 1.13 | IGFBP2 binds IGF-I and IGF-II in circulation and modulates IGF receptor signaling. IGFBP2 can indirectly influence immune responses and inflammation. | 17 |
| FGB | 1.13 | FGB encodes the beta chain of fibrinogen. During vascular injury, thrombin cleaves fibrinopeptides A and B from fibrinogen and converts it to fibrin for blood clot formation. | 18 |
| SNED1 | 1.08 | SNED1 is a secreted protein that localizes to the extracellular matrix and mediates cell-matrix adhesion. Breast cancer cells and neural crest cells have been shown to adhere to SNED1 through its RGD motif via integrins $\alpha 5\beta 1$ and $\alpha v\beta 3$. | 19 |
| APOH | 1.03 | APOH primarily functions as a coagulation factor and phospholipid-binding protein. It also acts as a scavenger of lipopolysaccharide (LPS) and certain pathogens. APOH is classified as an acute-phase protein and is upregulated during infection and inflammation. | 20 |

| | | | |
|---|---|---|---|
| JCHAIN | 1 | JCHAIN encodes the joining chain and promotes the assembly of IgA dimers and IgM pentamers. It enables binding to the polymeric immunoglobulin receptor (pIgR) on epithelial cells and facilitates trans-epithelial transport of secretory IgA and IgM into mucosal secretions. Expression of JCHAIN is largely restricted to B cells and plasma cells that produce polymeric IgA and IgM. | 21 |
| MENT | 0.99 | MENT is a member of the serpin (serine protease inhibitor) superfamily. It is a non-histone chromatin-associated protein that condenses chromatin into a more compact state and inhibits papain-family cysteine proteases. | 22 |
| F12 | 0.97 | F12 encodes the zymogen form of Factor XII. Once activated, Factor XIIa participates in the intrinsic coagulation cascade by activating Factor XI. | 23 |
| FGA | 0.97 | FGA encodes the alpha chain of fibrinogen which is a key coagulation protein | 24 |
| C1QTNF3-3 | 0.91 | C1QTNF3 (CTRP3) exerts anti-inflammatory and endothelial-protective effects. It dampens TLR4-driven NF-κB signaling, reduces endothelial expression of VCAM-1 and ICAM-1, decreases monocyte adhesion, and activates pro-survival PI3K/Akt/eNOS pathways. | 25, 26 |
| EFEMP1 | 0.88 | EFEMP1 encodes a secreted extracellular matrix glycoprotein. It interacts with tissue inhibitor of metalloproteinases-3 (TIMP-3) to modulate matrix metalloproteinase (MMP) activity and participates in cell-matrix | 27 |

| | | | |
|---|---|---|---|
| | | adhesion. | |
| CFHR5 | 0.85 | CFHR5 encodes a member of the Factor H-related (FHR) family of complement regulators. It binds complement C3 and is thought to modulate alternative pathway activity at sites of complement activation. | 28 |
| PCSK9 | 0.8 | PCSK9 is a regulator of cholesterol metabolism. It binds the low-density lipoprotein receptor (LDLR) and directs it for lysosomal degradation. Beyond lipid metabolism, PCSK9 can promote pro-inflammatory cytokine production and enhance NF-κB/TLR4-mediated signaling in macrophages and vascular cells. | 29, 30 |
| CRTAC1 | 0.78 | CRTAC1 encodes a cartilage-derived glycoprotein. It has been identified as a biomarker for osteoarthritis and lung adenocarcinoma. | 31 |
| MST1 | 0.77 | MST1 regulates T cell and B cell development and trafficking. It also participates in macrophage function, reactive oxygen species production, and inflammasome activation. | 32, 33 |
| CDH13 | 0.76 | CDH13 encodes a GPI-anchored cadherin that regulates GABAergic modulation in stem cell-derived neuronal networks. | 34 |
| IGFBP4 | 0.75 | IGFBP4 is a member of the insulin-like growth factor binding protein (IGFBP) family. It binds both IGF-I and IGF-II in circulation and modulates IGF bioavailability and receptor signaling | 35 |
| APMAP | 0.73 | APMAP localizes to the endoplasmic reticulum, where it suppresses lipid oxidation and supports lipoprotein processing. | 36 |

| | | | |
|---|---|---|---|
| | | These pathways can modulate innate immune responses in infected or inflamed tissues. | |
| CFD | 0.73 | CFD encodes a serine protease secreted primarily by adipocytes. It serves as a rate-limiting enzyme of the alternative complement pathway by cleaving factor B when complexed with C3b to generate the C3 convertase (C3bBb) and thus amplifies complement activation. | 37 |
| EN1 | 0.71 | EN1 has been linked to the fibroblast-to-myofibroblast transition and fibrotic disease. | 38 |
| HABP2 | 0.69 | HABP2 is a regulator of vascular endothelial barrier integrity. In models of acute lung injury, LPS stimulation increases HABP2 expression in pulmonary endothelial cells. Silencing HABP2 with small-interfering RNA attenuates LPS- and low-molecular-weight hyaluronan-induced endothelial hyperpermeability. HABP2 compromises vascular integrity through activation of a protease-activated receptor (PAR)-RhoA-Rho kinase signaling pathway. | 39 |
| FLT4 | 0.67 | FLT4 encodes a receptor tyrosine kinase that plays a role in lymphatic vessel formation and the maintenance of the lymphatic endothelium. | 40 |
| FGFR1 | 0.66 | FGFR1 encodes a receptor tyrosine kinase that binds fibroblast growth factor ligands and activates downstream signaling pathways that regulate cell proliferation and survival. | 41 |

| | | | |
|---|---|---|---|
| CFHR2 | 0.66 | CFHR2 is a regulator of the alternative complement pathway. It binds C3b and permits assembly of the C3 convertase but suppresses its catalytic activity and blocks formation of the terminal complement complex (TCC). | 42 |
| CD5L | 0.65 | CD5L encodes a soluble glycoprotein of the scavenger receptor cysteine-rich (SRCR) superfamily. It is predominantly produced and secreted by macrophages in lymphoid organs, liver, spleen, and inflamed tissues. Expression of CD5L is regulated by lipid-sensing nuclear receptors. | 43 |
| THBS4 | 0.65 | THBS4 encodes a thrombospondin family extracellular matrix glycoprotein. THBS4 participates in tissue remodeling and wound repair and contributes to vascular and ECM adaptive responses under stress. | 44 |
| COL6A1 | 0.64 | COL6A1 encodes the alpha-1 chain of type VI collagen which forms microfilament networks and contributes to cellular structural support. | 45 |
| ADAMTSL4 | 0.61 | ADAMTSL4 is a secreted protein that localizes to the extracellular matrix. It binds fibrillin-1 microfibrils and promotes microfibril assembly in tissue. | 46 |

| Down-Regulated Protein (Gene Name) | $Log_2$ Fold Change | Characterization of protein | Supplemental Citations |
|---|---|---|---|
| TNC | -0.59 | TNC is induced in tissues by pro-inflammatory stimuli and engages TLR4 to activate innate immune signaling and tissue | 47, 48 |

| | | | |
|---|---|---|---|
| | | repair. It modulates leukocyte behavior and contributes to inflammation during infection. | |
| DSG2 | -0.59 | DSG2 is essential for intestinal epithelial barrier integrity through its regulation of tight junction composition. Loss of DSG2 adhesion promotes inflammatory signaling and barrier dysfunction. | 49, 50, 51 |
| PRDX2 | -0.59 | PRDX2 functions as an intracellular antioxidant by catalyzing the reduction of hydrogen peroxide and organic hydroperoxides. Oxidized and glutathionylated PRDX2 can be released extracellularly and act as a damage-associated molecular pattern (DAMP). | 52 |
| SERPINA6 | -0.6 | SERPINA6 encodes corticosteroid-binding globulin (CBG) which is the primary plasma transport protein for glucocorticoids. It is produced mainly by the liver and circulates in plasma bound to cortisol. | 53 |
| CFB | -0.6 | CFB is a component of the alternative complement pathway. It enhances opsonization of pathogens through C3b deposition and facilitates phagocytosis. CFB also serves as a downstream effector of Toll-like receptor (TLR) signaling. Dysregulation of CFB has been implicated in complement-mediated diseases. | 54, 55 |
| HGFAC | -0.62 | HGFAC is primarily produced by hepatocytes and circulates in plasma as a zymogen. Its activation is associated to processes of tissue injury, coagulation and remodelling. | 56 |

| | | | |
|---|---|---|---|
| C1QA | -0.62 | C1QA encodes the A-chain polypeptide of complement subcomponent C1q. Together with the B and C chains, it forms the C1q molecule that binds antigen-antibody complexes and initiates cleavage of complement components C4 and C2. Mutations in C1QA are associated with autoimmunity and impaired clearance of apoptotic cells and immune complexes. | 57 |
| SERPINA3 | -0.63 | Elevated SERPINA3 levels have been associated with cerebral small-vessel disease severity. | 58 |
| GPX3 | -0.63 | GPX3 is a member of the glutathione peroxidase family. It is produced mainly by the kidneys and catalyzes the reduction of hydrogen peroxide and organic hydroperoxides using glutathione as a cofactor. | 59 |
| HRG | -0.65 | HRG encodes histidine-rich glycoprotein which is a plasma protein that modulate macrophage binding and coagulation. | 60 |
| SERPINA1 | -0.7 | SERPINA1 encodes alpha-1-antitrypsin which is a glycoprotein primarily synthesized by hepatocytes. It inhibits neutrophil elastase and other serine proteases to protect tissues from protease-mediated damage. | 61 |
| PDIA3 | -0.7 | PDIA3 encodes protein disulfide isomerase A3 which catalyzes disulfide bond formation through interactions with molecular chaperones. It plays a role in antigen presentation and is upregulated during infection. | 62 |

| | | | |
|---|---|---|---|
| SELL | -0.83 | SELL encodes L-selectin which is a cell-surface adhesion molecule of the selectin family expressed on most circulating leukocytes. It mediates the initial tethering and rolling of leukocytes on endothelial cells of high endothelial venules and inflamed microvessels. It facilitates leukocyte migration into tissues and lymphoid organs. | 63 |
| NRP1 | -0.85 | NRP1 encodes a transmembrane glycoprotein co-receptor expressed on neurons, endothelial cells, and various immune cell types. It contributes to immune regulation and cell migration. NRP1 has also been shown to regulate the stability and function of regulatory T cells. | 64 |
| ALDOA | -0.85 | ALDOA encodes aldolase A. | 65 |
| LDHA | -1.01 | LDHA encodes a subunit of lactate dehydrogenase. Elevated LDHA expression in tumors leads to lactate accumulation, which has been shown to suppress T cell function. | 66 |
| PRKCSH | -1.03 | PRKCSH encodes the beta subunit of glucosidase II which is an enzyme that resides in the endoplasmic reticulum and is involved in N-linked oligosaccharides and glycoprotein trafficking. In cancer, overexpression of PRKCSH has been associated with worse clinical outcomes. | 67 |
| ENPP2 | -1.03 | ENPP2 encodes a secreted lysophospholipase D that hydrolyzes lysophosphatidylcholine into lysophosphatidic acid (LPA). It plays a role in cell migration and has been associated with inflammation and cancer. | 68 |
